# Influence of Chinese eye exercises on myopia control in an East Asian population: a meta-analysis

**DOI:** 10.1101/19011270

**Authors:** Paradi Sangvatanakul, Jakkree Tangthianchaichana, Adis Tasanarong, Noel Pabalan, Phuntila Tharabenjasin

**Affiliations:** Chulabhorn International College of Medicine, Thammasat University, Pathum Thani, Thailand

**Keywords:** Chinese eye exercises, myopia, meta-analysis

## Abstract

**Objective:** The rates of myopia (−0.50 diopter), and high myopia (≥ −6.0 diopter) have been increasing in East Asian populations; the reasons for which may include the combinations of genetic, environment and behavioural factors. The most affected demography point to the young elite population of intellectuals produced from universities. Of the several recommendations to address the myopia epidemic, the influence of Chinese eye exercises (CEE) have been examined. However, reports have been inconsistent, prompting a meta-analysis to obtain more precise estimates.

**Methods:** Eight articles were included in the meta-analysis where we operated on the hypothesis that CEE either increased or reduced myopia control. We compared the subjects that performed CEE against those that did not. We used and estimated odds ratios [ORs] and 95% confidence intervals (CIs) using the generic inverse variance method. Subgroup analysis involved quality (high/serious and low/non-serious) and frequency (> 5 times/week) of performing CEE comprised. Heterogeneity was subjected to outlier treatment which split the findings into pre- (PRO) and post- (PSO) outlier. The strength of evidence in our findings were based on high significance (P^a^ < 10^−5^), surviving the Bonferroni correction and homogeneity (I^2^ = 0%). Outcomes with these features comprised our core findings.

**Results:** Our core findings were found in the PSO overall indicating elevated myopia control (OR 0.72, 95% CI 0.61-0.86, P^a^ = 0.0002) and CEE subgroups (Serious: OR 0.75, 95% CI 0.68-0.84, P^a^ < 10^−5^; Frequent: OR 0.55, 95% CI 0.45-0.68, P^a^ < 10^−5^). The low quality subgroup outcome was null in PRO (OR 0.97, 95% CI 0.50-1.86, P^a^ = 0.92) but conveyed significantly less myopia control in PSO (OR1.57, 95% CI 1.24-2.01, P^a^ = 0.0002).

**Conclusions:** This meta-analysis found that CEE afforded 28% greater control of myopia. Enabled by outlier treatment, this finding was homogeneous and consistent. Subgroup effects elevated myopia control to 62% when CEE was done up to 5 times a week. Improper CEE performance implied reduced myopia control of up to 57%.

## Introduction

Myopia is one of the most commonly occurring defects of the human eye refraction. Defined as ≥ −0.50 diopter (D), this ocular defect has three levels, low (≤−3.0 D), medium (−3.0 D to −5.9 D) and high (≥ −6.0 D). About one-fifth of the myopic global population can develop high myopia (≥ −6.0 D) rendering this eye disorder a significant epidemiological concern [1,2]. This concern is highlighted with a worldwide estimate of ∼ 1.4 billion myopic individuals comprising > 20% of the global population [3]. Moreover, this percentage is projected to increase to 50% (∼ 4.7 billion) by the year 2050 [3]. This increase in myopia prevalence will have substantial social, educational, and economic consequences to modern society. While geographic distribution of myopia prevalence varies between countries, this eye defect has become epidemic in East and Southeast Asia [3] with the fastest growing tendency pointing to the countries of Singapore and China [4]. Demographic comparisons among 12-year-olds show higher prevalence of myopia in Asians (Singapore: 62%; China: 49.7%) than Caucasians (USA: 20%; Australia: 11.9%) [5,3]. In a study of Chinese schoolchildren, myopia was reported to be present among 33.6% of first graders and 54% of seventh graders [6]. Yet, these percentages pale when compared with older Chinese teenagers and young adults where myopia is present in 90% of this demography [7]. Even more serious prevalence data was shown by a Shanghai study where myopia was present among 94.9% of undergraduate students and 96.9% of postgraduate students, and 19.5% of all myopic students had high myopia [8]. Data from the Ministry of Education of the People’s Republic of China in 2017 show that there are about 27.5 million university or college students affected with myopia [9]. Age-wise, myopia currently affects people from 10 to 39 years, but this demographic age range is predicted to widen from 10 to 79 years by the year 2050 [3]. Etiology of myopia is complex with genetic and environmental factors playing a role. Associations of genetic polymorphisms with myopia have been investigated [10-12], so has the impact of family history on the incidence of myopia among children [13,14]. Even high intelligence was shown to be associated with myopia [15]. Consequently, populations with a higher level of education are reported to have higher proportions of myopia [16]. Thus, we look at the intertwining of high educational attainment with genetics as influencing myopia prevalence [17]. The complication of environmental factors affecting myopia is that they are tied to human behavior, which in the modern context, is also tethered to the influence of technology (smartphones and computers). Still, these behavioral factors and technological influence afford greater human direct control compared to genetics in addressing the myopia epidemic. A major contributory factor in the environmental context point to the ubiquitous use of smartphones and computers [7], which dictates the behaviors of those at most risk for myopia. Risk behaviors for myopia include the following: One, high educational pressure in East and Southeast Asia does not permit children to spend extended time outdoors [18,19]. Thus, less outdoor activities promotes sedentary behavior. Two, long hours of exposure to digital screens [20]. Three, continuous reading without rests, has been shown to lead to myopia [21,22]. Myopia not only impacts upon physical health, but also increases risk of eye complications, including myopic retinopathy, myopic glaucoma, retinal detachments, and blindness [23-25]. High myopia (≥ −6.0 D) can result in loss of vision due to retinal detachment, neovascularization, cataract, glaucoma, or macular atrophy [1,26]. It can also cause impaired vision and blindness [27]. Interventions have been developed to prevent and control myopia [28]. Increased time for outdoor activities could effectively prevent myopia [29-32]. In its presence, however, myopia progression is hindered with low-dose atropine and orthokeratology [33,34]. Even then, these two therapeutic approaches may only be used for myopic control, rather than prevention. Therefore, there is a need to introduce a simple and easy-to-use intervention for myopia control. Chinese eye exercises (CEE), an intervention for visual protection and myopia prevention and progression, originated from the theories of Traditional Chinese Medicine [35]. Yet, reports of CEE effects in preventing myopia have been inconsistent where studies have suggested reduced risk and others have shown risk increases. This prompted a meta-analysis to obtain a more precise estimate of association. We aim to foster better understanding of the role of CEE in myopia onset and progression.

## Materials and Methods

### Selection of studies

We searched MEDLINE using PubMed, Google Scholar and Science Direct for relevant publications as of October 01, 2019. Terms used were “*Chinese eye exercises* “, and “*myopia*”, as medical subject heading and text, unrestricted by language. References cited in the retrieved articles were also screened manually to identify additional eligible studies. In cases of duplicate articles, we selected the one with a later date of publication. Inclusion criteria were (1) case–control design evaluating the association between CEE and myopia; (2) eye exercise in the presence and absence of myopia and (3) presence of raw frequency data or presence of odds ratios (ORs) and 95% confidence intervals (CIs). Exclusion criteria were (1) those not involving myopia and CEE; (2) reviews; and (3) studies whose raw data were unusable/absent. Primary study authors were contacted in order to obtain more information on incomplete data.

### Data extraction

Two investigators (PS and NP) independently extracted data. Disagrements were adjudicated by a third investigator (PT) until arrival at a consensus. The following information were obtained from each publication: first author’s name, published year, country of origin, age of participants, details of the myopia condition, and study design.

### Data distribution

Data distribution was assessed with the Shapiro-Wilks (SW) test using SPSS 20.0 (IBM Corp., Armonk, NY, USA). Normal distribution (P > 0.05) warranted descriptive and inferential expressions of mean ± standard deviation (SD) and the parametric approach, respectively. Otherwise, the median with interquartile range (IQR) and non-parametric tests were used, respectively.

### Methodological quality of the studies

We assessed the methodological quality of the included studies using two scales that were appropriate to the study design. The Jadad scale [36], for randomized control trials (RCTs) has five questions that examines randomization, double-blind and attrition. It provides a total score that ranges from zero to five, where zero indicates low quality and five, high quality [36]. We used the Newcastle-Ottawa Scale (NOS) for cross-sectional (xs) and case-control (cc) studies [37]. The ⋆- based NOS contains eight items with three dimensions (number of ⋆in parentheses): selection (4), comparability (1), and outcome (3). The NOS scores range from zero up to nine ⋆for cc studies and zero to 10 ⋆for xs studies. Low, moderate and high have scores of < 4, 4-5 and ≥ 6-7, respectively.

### Risk of bias

Risk of bias assessment followed the Cochrane handbook [38], which was applied to cc and RCT study designs [39]. This method evaluates biases originating from sequence generation (selection bias), allocation sequence concealment (selection bias), blinding of participants and personnel (performance bias), blinding of outcome assessment (detection bias), incomplete outcome data (attrition bias), and selective outcome reporting (reporting bias). Every item was judged as either yes, no, or unclear. More items judged as “yes” indicated a low likelihood of bias. In contrast, high likelihood of bias garnered more “no” judgements. Insufficient descriptions merited “unclear” judgment. Judgments were assigned by two of the authors (NP and PT) working independently, and discrepancies were remedied through discussions with a third investigator (PS) to obtain a consensus.

### Meta-analysis

Because we addressed the issues of myopia onset and progression, we hypothesized that performing CEE would afford greater or lesser control of myopia. Greater and lesser control entailed less and more risk, respectively. In order to quantify this control, we assessed the types of data provided by each article, which were ORs and 95% CIs or raw frequency data (number of participants who performed CEE versus those that did not). In the presence of both, we opted to extract the former. Presence of raw data prompted their extraction from each study and were used to calculate study-specific ORs with 95% CIs. The study-specific ORs were transformed logarithmically (log OR). We also derived the standard error (SE) from the 95% CI [40]. Using the generic inverse variant (GIV) method [41], log OR and SE from each paper were entered as the operating data in Review Manager (RevMan) 5.3 (Cochrane Collaboration, Oxford, England). Subgroup analysis consisted of high quality (HQ)/serious, low quality (LQ) and frequency of performing CEE. Heterogeneity between studies was estimated with the χ^2^-based Q test [42], with threshold of significance set at P^b^ < 0.10. Heterogeneity was also quantified with the I^2^ statistic which measures variability between studies [43]. I^*2*^ values of > 75% indicate substantial variability and 0%, zero heterogeneity (homogeneity). Evidence of functional similarities in population features of the studies warranted using the fixed-effects model [44], otherwise the random-effects model [41] was used. Sources of heterogeneity were detected with the Galbraith plot [45] followed by re-analysis (outlier treatment). Of note, outlier treatment dichotomized the comparisons into pre-outlier (PRO) and post-outlier (PSO). Sensitivity analysis, which involves omitting one study at a time and recalculating the pooled OR, was used to test for robustness of the summary effects. Publication bias was considered for assessment if the comparisons met two conditions: (i) ≥ 10 studies only [46] and (ii) significant outcomes. Multiple comparisons were Bonferroni-corrected. Except for heterogeneity estimation [42], two-sided P-values of ≤ 0.05 were considered significant. Other data were analyzed using SIGMASTAT 2.03, SIGMAPLOT 11.0 (Systat Software, San Jose, CA, USA) and SPSS 20.0 (IBM Co., Armonk, NY, USA).

## Results

### Search results and study features

Figure 1 outlines the study selection process in a PRISMA-sanctioned flowchart (Preferred Reporting Items for Systematic Reviews and Meta-Analyses). Initial search resulted in 60 citations, followed by a series of omissions that eventually yielded eight articles for inclusion [47-54].

**Fig 1.**
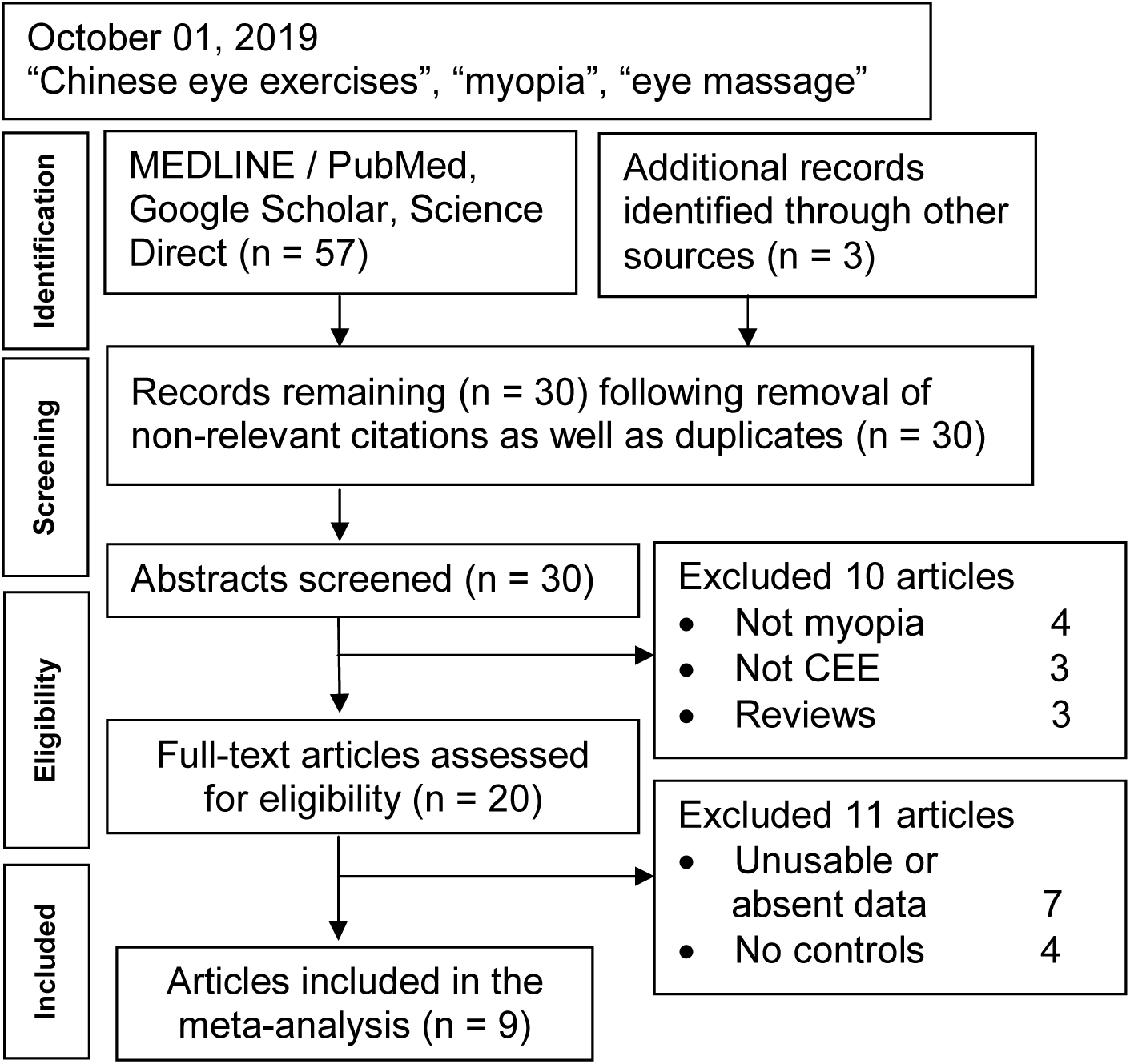
Summary flowchart of literature search.

### Characteristics of the included studies

Table 1 shows the year span of the included articles (2011-2019) and ethnic composition, Chinese (n = 6) and Indian (n = 2). Non-normal (SW: P = 0.022) age distribution of the participants in the collection of studies showed a young demography (median: 12.7 years, IQR: 11.3-20.7). Methodological quality of the component studies was generally high for xs but probably not for RCT studies. S1 and S2 Tables detail the scores for each study based on the Jadad (RCT) and NOS (cc/xs) scales. This meta-analysis followed the PRISMA guidelines (S3 Table).

**Table 1.** Characteristics of the included studies

| K | Lu et al | First author | [R] | Year | Country | Age (y) mean ± SD (range) | Myopia details | Study design | MQJ |
| --- | --- | --- | --- | --- | --- | --- | --- | --- | --- |
| 1 | ○ | Rathod | [47] | 2011 | India | 21 ± 1.4 | 30 subjects | RCT * | Poor |
| 2 | ✓ | Kang | [48] | 2012 | China | 12.7 ± 0.5 | School age | cc ** | High |
| 3 | ✓ | Lin | [49] | 2013 | China | 11.1 ± 3.2 | Acupoints urban | xs ** | High |
| 4 | ✓ | Han | [50] | 2014 | China | High school | High altitude | xs ** | Low |
| 5 | ○ | Li | [51] | 2015 | India | 10-14 | School age | RCT * | Good |
| 6 | ✓ | Lin | [52] | 2016 | China | 11.1 ± 2.4 | Acupoints rural | xs ** | High |
| 7 | ✓ | Wang | [53] | 2017 | China | 21.1 ± 1.6 | Medical students | xs ** | High |
| 8 | ○ | Huang | [54] | 2019 | China | 19.6 ± 0.9 | University | xs ** | High |

K: number designation of the article; (R) Reference; inclusion of the article in the previous meta-analysis (Lu et al 2019) is indicated by ✓, otherwise ○. SD: standard deviation; RCT: randomized control trial; cc: case-control; xs: cross-sectional; \* Jadad scale (S1 Table); \*\* NOS (S2 Table); MQJ: methodological quality judgment

### Risk of bias outcomes

Although Fig 2 shows a general low risk of bias based on the six criteria, Fig 3 delineates the poor quality of the Rathod et al study [47] which contrasted with the high quality of Li et al [51] and Kang et al [48].

**Fig 2.**
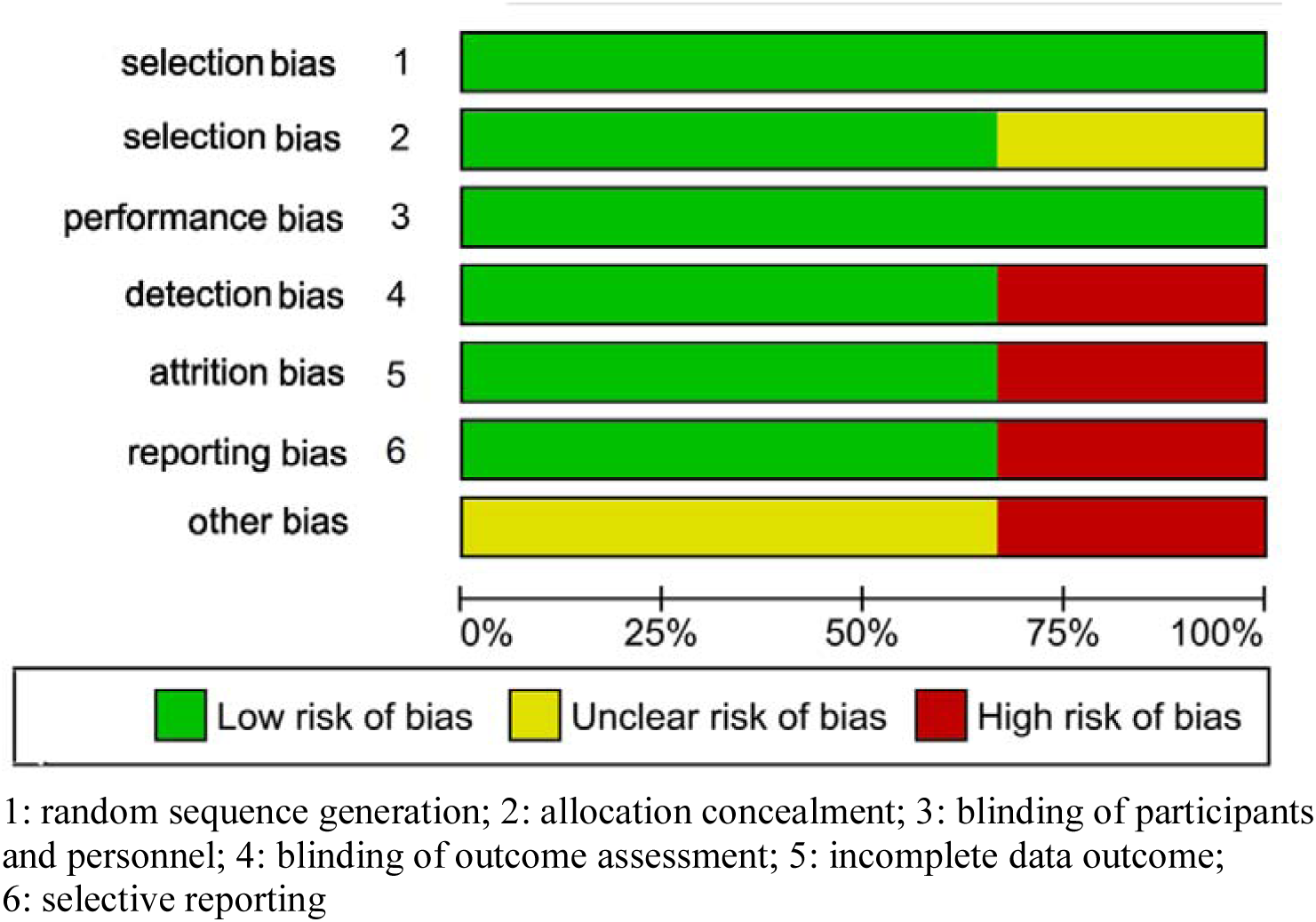
Risk of bias analysis of CEE influence on myopia using the Cochrane Collaboration tool 1: random sequence generation; 2: allocation concealment; 3: blinding of participants and personnel; 4: blinding of outcome assessment; 5: incomplete data outcome; 6: selective reporting

**Fig 3.**
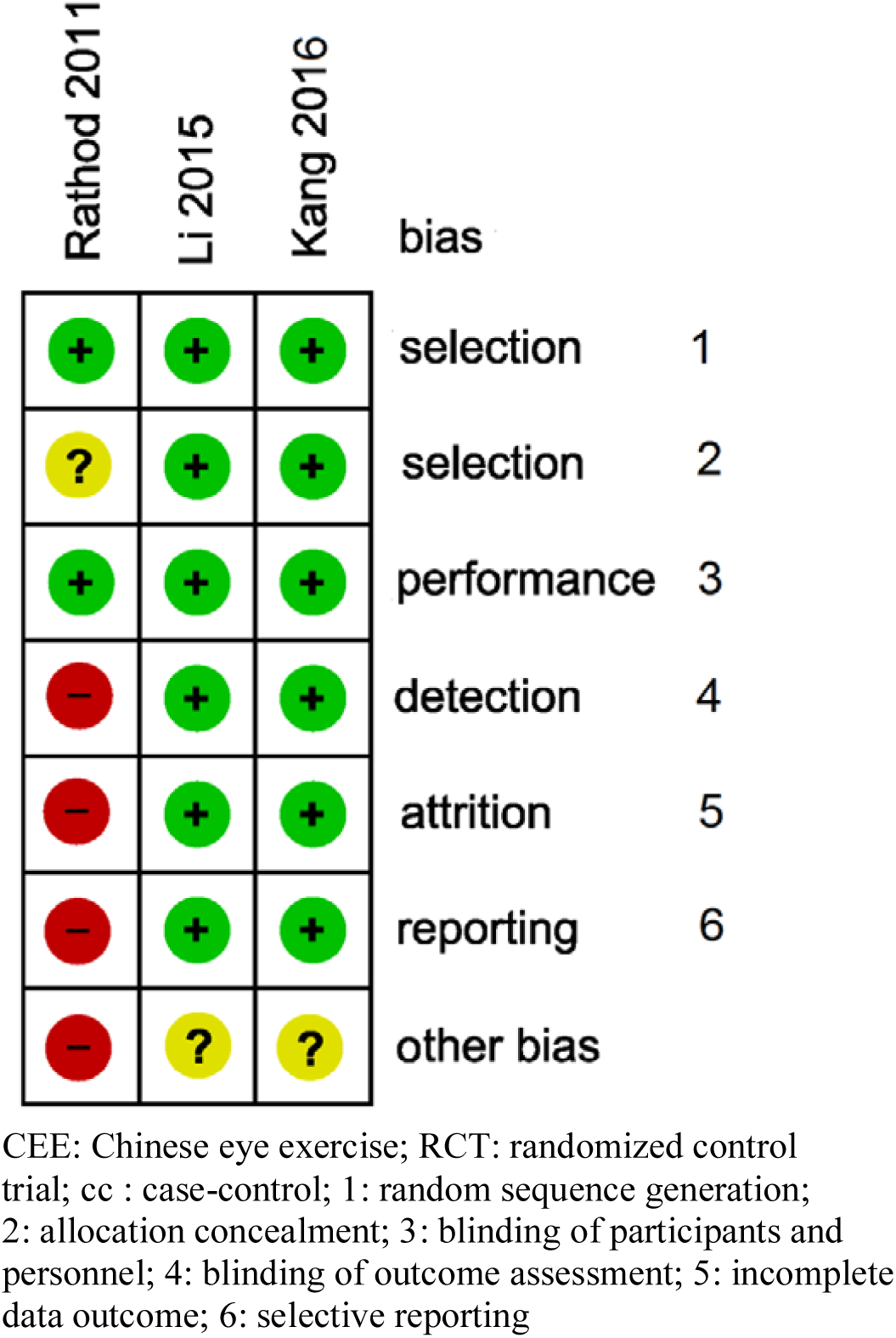
Risk of bias summary among and RCT/cc studies that examined the influence of CEE on myopia CEE: Chinese eye exercise; RCT: randomized control trial; cc: case-control; 1: random sequence generation; 2: allocation concealment; 3: blinding of participants and personnel; 4: blinding of outcome assessment; 5: incomplete data outcome; 6: selective reporting

### Meta-analysis outcomes

Table 2 shows six significant outcomes, four of which are attributed to outlier treatment. One finding indicating less myopia control was found in the LQ subgroup (OR 1.57, 95% CI 1.24-2.01, P^a^ = 0.0002). Otherwise, most of the findings indicate greater control of myopia. In PRO, OR magnitude was greater (ORs 0.38-0.49, 95% CIs 0.22-0.82, P^a^ = 0.0007-0.007) than PSO, whose findings are modulated (ORs 0.55-0.75, 95% CIs 0.45-0.86, P^a^ < 10^−5^-0.0002). Significance, however, was greater in PSO (up to P^a^ < 10^−5^) than in PRO (up to P^a^ = 0.0007).

**Table 2.** Summary associations between CEE and myopia

|  | Test of association |  |  |  | Test of heterogeneity |  |  |  | Test of association |  |  |  | Test of heterogeneity |  |  |  | Effect of outlier treatment |  |
| --- | --- | --- | --- | --- | --- | --- | --- | --- | --- | --- | --- | --- | --- | --- | --- | --- | --- | --- |
|  | n | OR | 95% CI | P<sup>a</sup> | MC | P<sup>b</sup> | I<sup>2</sup> (%) | AM | n | OR | 95% CI | P<sup>a</sup> | MC | P<sup>b</sup> | I<sup>2</sup> (%) | AM | Sig | Het |
|  | PRO |  |  |  |  |  |  |  | PSO |  |  |  |  |  |  |  |  |  |
| CEE | 9 | 0.84 | 0.48-1.49 | 0.55 | more | 10<sup>-5</sup> | 94 | R | 6 | **0.72** | **0.61-0.86** | **0.0002** ✓* | more | 0.95 | 0 | F | GS | EH |
| Serious (HQ) | 7 | **0.49** | **0.29-0.82** | **0.007** ✕* | more | 10<sup>-5</sup> | 97 | R | 4 | **0.75** | **0.68-0.84** | **10<sup>-5</sup>** ✓* | more | 0.46 | 0 | F | ES | EH |
| LQ | 6 | 0.97 | 0.50-1.86 | 0.92 | null | 10<sup>-5</sup> | 88 | R | 4 | **1.57** | **1.24-2.01** | **0.0002** ✓<sub>δ</sub> | less | 0.53 | 0 | F | GS | EH |
| Frequent | 5 | **0.38** | **0.22-0.67** | **0.0007** ✓* | more | 10<sup>-5</sup> | 95 | R | 3 | **0.55** | **0.45-0.68** | **10<sup>-5</sup>** ✓* | more | 0.12 | 52 | F | ES | RH |

CEE: Chinese eye exercise; HQ: high quality; LQ: low quality; n: number of studies; OR: odds ratio; CI: confidence interval; P<sup>a</sup>: P-value for association; in: increased risk; MC: myopia control; null: ORs 0.97-1.03; P<sup>b</sup>: P-value for heterogeneity; I<sup>2</sup>: measure of variability; AM: analysis model; R: random-effects; F: fixed-effects; PRO: pre-outlier; PSO: post outlier; RS: retained significance; RNS: retained non-significance; RH: reduced heterogeneity; EH: eliminated heterogeneity; values in bold indicate significant associations; Bonferroni correction (✕ did not survive, ✓ survived); \* robust; <sub>δ</sub> non-robust (Table 3).

### Mechanism of outlier treatment

The mechanism of outlier treatment for the CEE analysis is visualized in Figs 4-6. Fig 4 shows the PRO forest plot, non-significant (OR 0.84, 95% CI 0.48-1.49, P^a^ = 0.55) and heterogeneous (P^b^ < 10^−5^, I^2^ = 94%). The Galbraith plot identifies five studies in three articles [50-52] as the sources of heterogeneity (outliers), located above and below the +2 and −2 confidence limits (Fig 5). In Fig 6, the PSO outcome (outliers omitted) shows eliminated heterogeneity (P^b^ = 0.95, I^2^ = 0%); greater myopia control (OR 0.72, 95% CI 0.61-0.86) and escalated significance (P^a^ = 0.0002). This operation is numerically summarized in Table 2.

**Fig 4.**
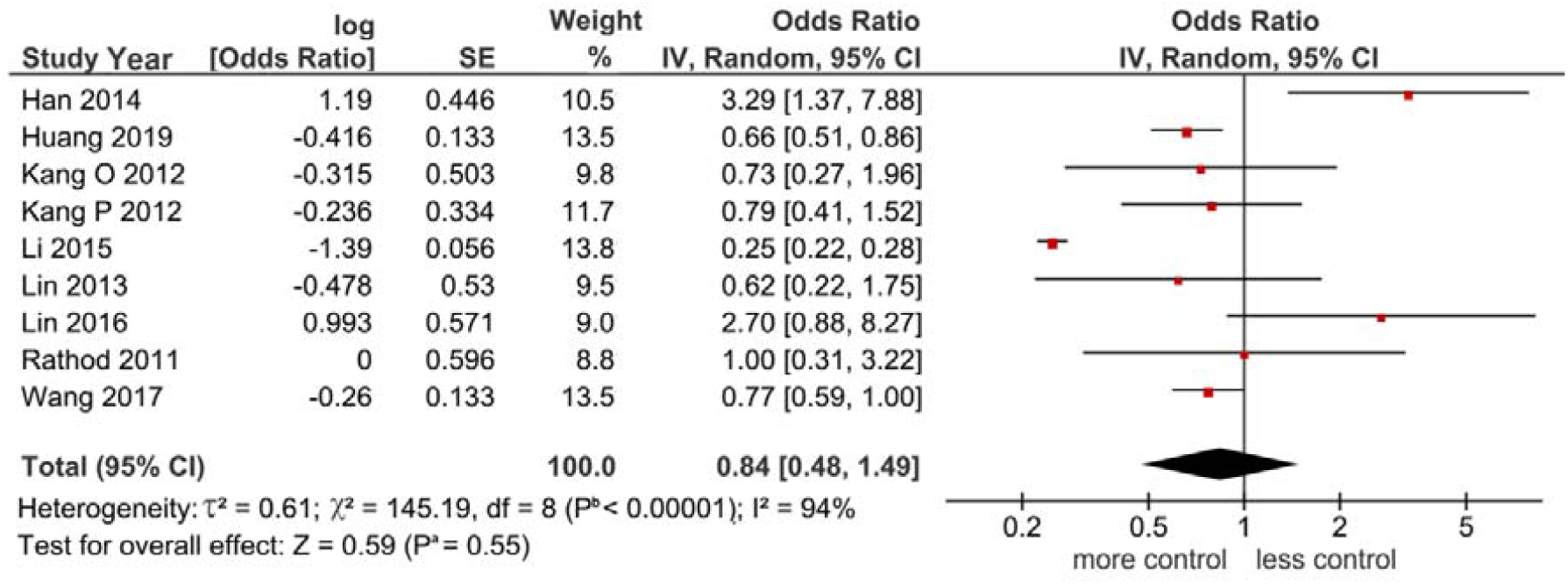
Forest plot outcome in the CEE analysis O: onset; P: progression; Diamond denotes the pooled odds ratio (OR), here indicating more myopia control (0.84). Squares indicate the OR in each study. Horizontal lines on either side of each square represent the 95% confidence intervals (CI). The Z test for overall effect is non-significant (P^a^ = 0.55). The chi-square test shows the presence of heterogeneity (P^b^ < 10^−5^, I^2^ = 94%); I^2^: a measure of variability expressed in %

**Fig 5.**
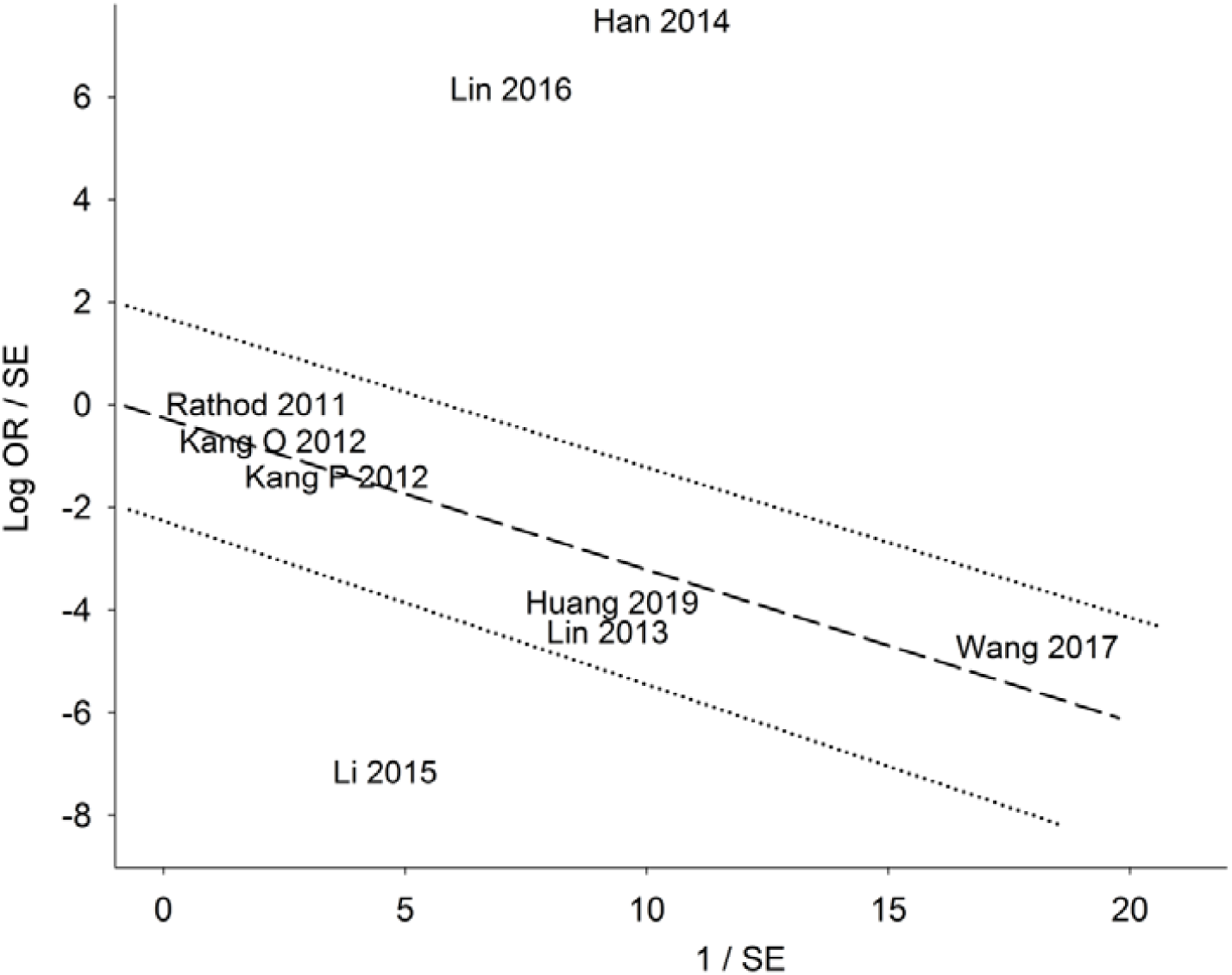
Galbraith plot in the CEE analysis log OR: logarithm of standardized odds ratio; SE: standard error. The two studies above the +2 and the one study below the −2 confidence limits are the outliers.

**Fig 6.**
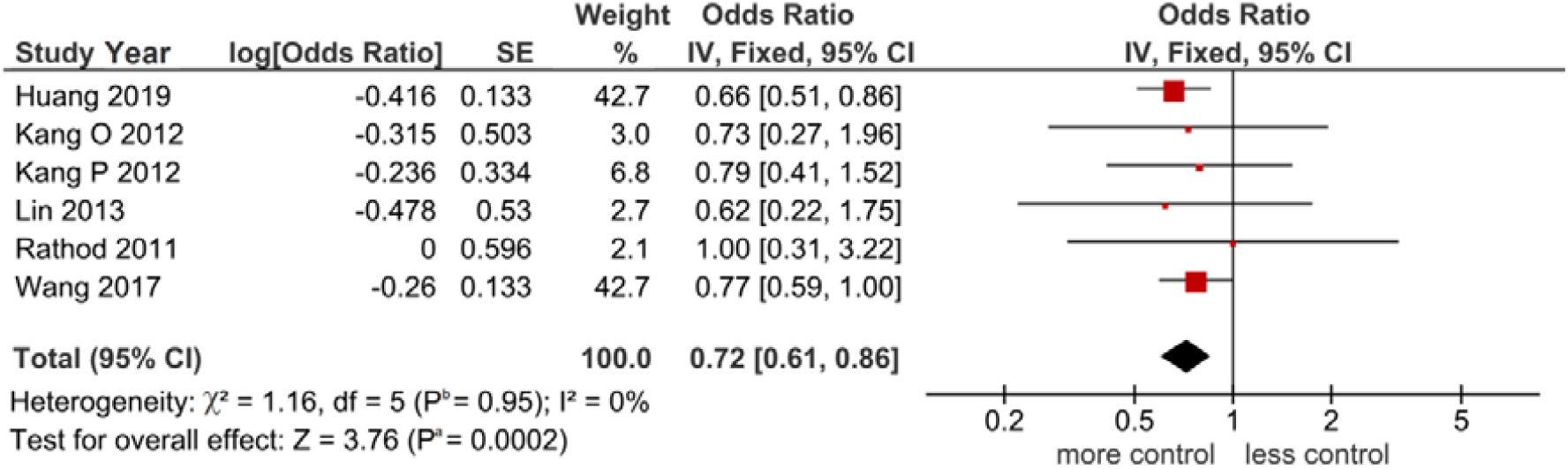
Forest plot outcome of outlier treatment in the CEE analysis O: onset; P: progression; The diamond denotes the pooled odds ratio (OR) indicating even more myopia control (0.72) compared with the forest plot in Fig 4. Squares indicate the OR in each study. Horizontal lines on either side of each square represent the 95% confidence intervals (CI). The Z test for overall effect was significant (P^a^ = 0.0002).The chi-square test indicates zero heterogeneity (P^b^ = 0.95, I^2^ = 0%); I^2^: a measure of variability expressed in %

### Sensitivity analysis and publication bias

Table 3 shows that five of the six significant outcomes were robust. Only the LQ comparison was non-robust on account of one study contributing to its stability [50]. We found no evidence of publication bias (Table 4).

**Table 3.** Outcomes of sensitivity treatment on the significant findings

| Comparison | OS | P<sup>a</sup> | Sensitivity outcome |
| --- | --- | --- | --- |
| CEE | PSO | 0.0002 | Robust |
| Serious (HQ) | PRO | 0.007 | Robust |
| Serious (HQ) | PSO | 10<sup>-5</sup> | Robust |
| Frequent | PRO | 0.0007 | Robust |
| Frequent | PSO | 10<sup>-5</sup> | Robust |
| LQ | PSO | 0.0002 | Han [50] |

CEE: Chinese eye exercise; OS: outlier status; HQ: high quality; LQ: low quality; PSO: post-outlier; PRO: pre-outlier; P<sup>a</sup>: P-value for association; bracketed number indicates the reference number, the study that contributed to non-robustness of the comparison

**Table 4.** Outcomes of publication bias tests on the significant findings

| Comparison | OS | P<sup>a</sup> | SW<br>P-value | Normal<br>distribution | PBT | Coefficient | PBT<br>P-value | Evidence<br>of PB |
| --- | --- | --- | --- | --- | --- | --- | --- | --- |
| CEE | PSO | 0.0002 | 0.421 | Yes | Egger | Intercept | 0.586 | None |
| Serious (HQ) | PRO | 0.007 | 0.316 | Yes | Egger | Intercept | 0.924 | None |
| Serious (HQ) | PSO | 10<sup>-5</sup> | 0.216 | Yes | Egger | Intercept | 0.140 | None |
| Frequent | PRO | 0.0007 | 0.092 | Yes | Egger | Intercept | 0.184 | None |
| Frequent | PSO | 10<sup>-5</sup> | 0.593 | Yes | Egger | Intercept | 0.999 | None |
| LQ | PSO | 0.0002 | 0.129 | Yes | Egger | Intercept | 0.943 | None |

CEE: HQ: high quality; LQ: low quality; OS: outlier status; P<sup>a</sup>: P-value for association; SW: Shapiro-Wilk; PBT: publication bias test

## Discussion

### Summary of associations

Our principal findings point to the consistency of significant control of myopia which is supported by homogeneity, robustness and stability. Outlier treatment has unraveled significant and homogeneous associations which were not present in the component single-study outcomes. Conflicting outcomes between primary studies may be attributed to small sample sizes. Outcomes based on small sample sizes appear to be common in clinical studies [55] and are prone to the risk of Type 1 error. Outlier treatment and subgroup analysis underpinned the importance of doing CEE properly and frequently (> 5 time/week) for better myopia control and highlighted the adverse consequence of improper CEE performance. This meta-analysis showed that outcomes were more dependent on xs than cc or RCTs. The xs studies had high sample sizes but not RCTs in our meta-analysis. This is unsurprising since RCTs require greater logistical organization than xs studies, which are mainly questionnaire-based. In fact, two xs studies [52,49] included in our meta-analysis recommended RCTs for better understanding of CEE influence on myopia.

### Comparison with a 2019 meta-analysis

We compare our findings with a recent meta-analysis (Lu et al. 2019) in terms of methodology, findings and conclusion [56]. The initial difference is that Lu et al examined myopia onset only, while we addressed both myopia onset and progression. Despite this difference, both meta-analyses used the GIV approach, lending comparability to the outcomes. Lu et al overall findings indicated increased risk for myopia onset (1.2-fold, n = 4) which contrasted with ours (28% more control of myopia, n = 6). Lu et al stated the harmful effects of low quality CEE on myopia which prompted us meta-analyze the data on low quality CEE from our included studies. Our initial low quality subgroup finding showed a null effect (OR 0.97, 95% CI 0.50-1.86, P = 0.92) until it was subjected to outlier treatment, the outcome of which indicated significantly less myopia control (OR 1.57, 95% CI 1.24-2.01, P = 0.0002). Thus, our PSO low quality finding concurred with the previous meta-analysis [56].

### CEE and myopia

CEE is an eye-care program that aims to protect against or prevent myopia and is mandated in Chinese schools [51]. Massaging the acupoints around the eyes accelerates the blood circulation, improves metabolism, rests the eye muscles, and relieves eye fatigue [57]. From the perspective of traditional Chinese Medicine, Qi is stimulated in meridians by massaging the acupoints, thus relieving eye strain [48]. By far, performing CEE was found to have a modest protective effect against myopia [52]. In contrast, non-performance of CEE was found to increase the risk for myopia [53]. A third variation in CEE study findings was the non-significant association between performing CEE and risk of myopia onset and myopia progression [48]. These associative variations may be due to the limited sample sizes and lack of rigor in measuring the quality of CEE performed. In schools, most Chinese children were required to perform CEE only once or twice a day, once in the morning and once in the afternoon, with 5 minutes for each time. Therefore, the total time of performing CEE was at most 10 minutes a day and in some places the children only performed the exercise once a day [48]. Studies have reported that most children could not perform the CEE with standard manipulation. Previous studies showed that about 90% of Chinese children did not perform CEE correctly, most of them could not find the exact periocular acupoints and did not have accurate pressure and manipulation skills for the exercises although they did them every day [58,59]. Our findings pointed to a significant 57% reduced control of myopia, which may exacerbate the coonidtion when performed improperly. On the other hand, we also showed significantly better control (up to 51%) of myopia (which may retard its progression or reduce the onset) when CEE is performed seriously. About 15% of total children and about a third of those who performed them were found to achieve high quality CEE [48].

### Strengths and limitations

Interpreting our findings should consider its limitations and strengths. Limitations include: (i) Given the greater reliance on RCTs over xs studies [52,49] limits our meta-analysis findings because xs data preclude direct evidence on the association between CEE and myopia [52]. (ii) All the component studies were underpowered; (iii) the low quality subgroup finding was non-robust; (iii) although less prevalent, myopia is not limited to the populations studied here, thus rendering the results inapplicable to other ethnic groups. (v) Clinically, the long-term effect of myopia prevention or slowing its progression remains an open question [51]. (vi) The likelihood of recall bias regarding responses of the participants to the questionnaires may have affected the accuracy of our study. However, impact of these limitations may have been mitigated with our assessments of the risk of bias and methodological quality of the studies. On the other hand, the strengths comprise of the following: (i) our use of the GIV allowed comparison with a previous meta-analysis. (ii) Outlier treatment was key to generating significance and eliminating heterogeneity which underpinned our core findings. This demonstrates the utility of this meta-analysis tool in elevating the level of evidence for associations; (iii) subgroup analysis validated the overall findings, allowed comparison and confirmed the adverse effect of LQ performance of CEE; (iv) all significant PSO ORs survived the Bonferroni correction, thus minimizing the possibility of a Type 1 error; (v) sensitivity treatment conferred robustness to most of the significant findings; (vi) no evidence of publication bias.

## Conclusions

We have shown the significant influence of CEE in controlling myopia, provided the performance was serious and frequent. These findings are buttressed with layers of evidential strength which include high statistical significance, stability and homogeneity.The substantial amount of evidence presented here may render CEE as a useful preventative and therapeutic option in myopia control. In spite of the evidence for associations, the complexity of CEE performance involves interactions between genetic and non-genetic factors that may not have been covered in this meta-analysis because of the logistical limitations. Additional well-designed studies exploring other parameters would confirm or modify our results and add to the extant knowledge about the influence of of CEE in controlling myopia

## Data Availability

Available upon request

## List of abbreviations

AM: analysis model
cc: case-control
CEE: Chinese eye exercises
CI: confidence interval
D: diopter
EH: eliminated heterogeneity
F: fixed-effects
GS: gained significance
GIV: generic inverse variance
I^2^: measure of variability
IQR: interquartile range
Log: logarithm
LS: lost significance
maf: minor allele frequency
MC: myopia control
MQJ: methodological quality judgment
n: number of studies
NOS: Newcastle-Ottawa Scale
OR: odds ratio
P^a^: P-value for association
P^b^: P-value for heterogeneity
PRISMA: Preferred Reporting Items for Systematic Reviews and Meta-Analyses
PRO: pre-outlier
PSO: post-outlier
R: random-effects
[R]: Reference
RCT: randomized control trial
RNS: retained non-significance
RH: reduced heterogeneity
SD: standard deviation
SE: standard error
SW: Shapiro-Wilk
xs: cross-section

## Data availability

The data [frequency, odds ratios and 95% confidence intervals] supporting this meta-analysis are from previously reported studies and datasets, which have been cited. The processed data are available from the corresponding author upon request.

## Supplementary materials

S1 Table Jadad DOCX

S2 Table NOS DOCX

S3 Table PRISMA DOCX

## Author contributions

**Conceptualization:** PS, JT

**Formal analysis:** NP, PT, PS

**Investigation:** NP, PT, PS

**Methodology:** NP, PT

**Resources:** PS, JT, AT

**Supervision:** AT

**Validation:** NP, PT, KP

**Writing – original draft:** NP, PT, PS

**Writing – review & editing:** NP, PT, PS, AT

## References

1. Morgan IG, French AN, Ashby RS, Guo X, Ding X, He M, Rose KA (2018) The epidemics of myopia: Aetiology and prevention. Progress in retinal and eye researchx 62:134–149. doi:10.1016/j.preteyeres.2017.09.004

2. Morgan IG, Ohno-Matsui K, Saw SM (2012) Myopia. Lancet 379 (9827):1739–1748. doi:10.1016/S0140-6736(12)60272-4

3. Holden BA, Fricke TR, Wilson DA, Jong M, Naidoo KS, Sankaridurg P, Wong TY, Naduvilath TJ, Resnikoff S (2016) Global Prevalence of Myopia and High Myopia and Temporal Trends from 2000 through 2050. Ophthalmology 123 (5):1036–1042. doi:10.1016/j.ophtha.2016.01.006

4. Ding BY, Shih YF, Lin LLK, Hsiao CK, Wang IJ (2017) Myopia among schoolchildren in East Asia and Singapore. Survey of ophthalmology 62 (5):677–697. doi:10.1016/j.survophthal.2017.03.006

5. Foster PJ, Jiang Y (2014) Epidemiology of myopia. Eye 28 (2):202–208. doi:10.1038/eye.2013.280

6. Li L, Zhong H, Li J, Li CR, Pan CW (2018) Incidence of myopia and biometric characteristics of premyopic eyes among Chinese children and adolescents. BMC ophthalmology 18 (1):178. doi:10.1186/s12886-018-0836-9

7. Dolgin E (2015) The myopia boom. Nature 519 (7543):276–278. doi:10.1038/519276a

8. Sun J, Zhou J, Zhao P, Lian J, Zhu H, Zhou Y, Sun Y, Wang Y, Zhao L, Wei Y, Wang L, Cun B, Ge S, Fan X (2012) High prevalence of myopia and high myopia in 5060 Chinese university students in Shanghai. Investigative ophthalmology & visual science 53 (12):7504–7509. doi:10.1167/iovs.11-8343

9. PRC MoE (2019) Number of Students of Formal Education by Type and Level 2018 http://www.moe.gov.cn/s78/A03/moe_560/jytjsj_2017/qg/201808/t20180808_344698.html Accessed 01 October 2019. Accessed 01 October 2019

10. Gong B, Qu C, Huang XF, Ye ZM, Zhang DD, Shi Y, Chen R, Liu YP, Shuai P (2016) Genetic association of COL1A1 polymorphisms with high myopia in Asian population: a Meta-analysis. International journal of ophthalmology 9 (8):1187–1193. doi:10.18240/ijo.2016.08.16

11. Tang SM, Lau T, Rong SS, Yazar S, Chen LJ, Mackey DA, Lucas RM, Pang CP, Yam JC (2019) Vitamin D and its pathway genes in myopia: systematic review and meta-analysis. The British journal of ophthalmology 103 (1):8–17. doi:10.1136/bjophthalmol-2018-312159

12. Zhang D, Zeng G, Hu J, McCormick K, Shi Y, Gong B (2017) Association of IGF1 polymorphism rs6214 with high myopia: A systematic review and meta-analysis. Ophthalmic genetics 38 (5):434–439. doi:10.1080/13816810.2016.1253105

13. Xiang F, He M, Morgan IG (2012) The impact of severity of parental myopia on myopia in Chinese children. Optometry and vision science: official publication of the American Academy of Optometry 89 (6):884–891. doi:10.1097/OPX.0b013e318255dc33

14. Mutti DO, Mitchell GL, Moeschberger ML, Jones LA, Zadnik K (2002) Parental myopia, near work, school achievement, and children’s refractive error. Investigative ophthalmology & visual science 43 (12):3633–3640

15. Verma A, Verma A (2015) A novel review of the evidence linking myopia and high intelligence. Journal of ophthalmology 2015:271746. doi:10.1155/2015/271746

16. Mirshahi A, Ponto KA, Hoehn R, Zwiener I, Zeller T, Lackner K, Beutel ME, Pfeiffer N (2014) Myopia and level of education: results from the Gutenberg Health Study. Ophthalmology 121 (10):2047–2052. doi:10.1016/j.ophtha.2014.04.017

17. Verhoeven VJ, Buitendijk GH, Consortium for Refractive E, Myopia, Rivadeneira F, Uitterlinden AG, Vingerling JR, Hofman A, Klaver CC (2013) Education influences the role of genetics in myopia. European journal of epidemiology 28 (12):973–980. doi:10.1007/s10654-013-9856-1

18. Zhou Z, Morgan IG, Chen Q, Jin L, He M, Congdon N (2015) Disordered sleep and myopia risk among Chinese children. PloS one 10 (3):e0121796. doi:10.1371/journal.pone.0121796

19. Morgan IG, French AN, Rose KA (2018) Intense schooling linked to myopia. Bmj 361:k2248. doi:10.1136/bmj.k2248

20. Fernandez-Montero A, Olmo-Jimenez JM, Olmo N, Bes-Rastrollo M, Moreno-Galarraga L, Moreno-Montanes J, Martinez-Gonzalez MA (2015) The impact of computer use in myopia progression: a cohort study in Spain. Preventive medicine 71:67–71. doi:10.1016/j.ypmed.2014.12.005

21. Ip JM, Saw SM, Rose KA, Morgan IG, Kifley A, Wang JJ, Mitchell P (2008) Role of near work in myopia: findings in a sample of Australian school children. Investigative ophthalmology & visual science 49 (7):2903–2910. doi:10.1167/iovs.07-0804

22. You X, Wang L, Tan H, He X, Qu X, Shi H, Zhu J, Zou H (2016) Near Work Related Behaviors Associated with Myopic Shifts among Primary School Students in the Jiading District of Shanghai: A School-Based One-Year Cohort Study. PloS one 11 (5):e0154671. doi:10.1371/journal.pone.0154671

23. Liu HH, Xu L, Wang YX, Wang S, You QS, Jonas JB (2010) Prevalence and progression of myopic retinopathy in Chinese adults: the Beijing Eye Study. Ophthalmology 117 (9):1763–1768. doi:10.1016/j.ophtha.2010.01.020

24. Saw SM, Gazzard G, Shih-Yen EC, Chua WH (2005) Myopia and associated pathological complications. Ophthalmic & physiological optics: the journal of the British College of Ophthalmic Opticians 25 (5):381–391. doi:10.1111/j.1475-1313.2005.00298.x

25. Xu L, Wang Y, Wang S, Wang Y, Jonas JB (2007) High myopia and glaucoma susceptibility the Beijing Eye Study. Ophthalmology 114 (2):216–220. doi:10.1016/j.ophtha.2006.06.050

26. Zejmo M, Forminska-Kapuscik M, Pieczara E, Filipek E, Mrukwa-Kominek E, Samochowiec-Donocik E, Leszczynski R, Smuzynska M (2009) Etiopathogenesis and management of high-degree myopia. Part I. Medical science monitor: international medical journal of experimental and clinical research 15 (9):RA199–202

27. Wong TY, Ferreira A, Hughes R, Carter G, Mitchell P (2014) Epidemiology and disease burden of pathologic myopia and myopic choroidal neovascularization: an evidence-based systematic review. American journal of ophthalmology 157 (1):9–25 e12. doi:10.1016/j.ajo.2013.08.010

28. Huang J, Wen D, Wang Q, McAlinden C, Flitcroft I, Chen H, Saw SM, Chen H, Bao F, Zhao Y, Hu L, Li X, Gao R, Lu W, Du Y, Jinag Z, Yu A, Lian H, Jiang Q, Yu Y, Qu J (2016) Efficacy Comparison of 16 Interventions for Myopia Control in Children: A Network Meta-analysis. Ophthalmology 123 (4):697–708. doi:10.1016/j.ophtha.2015.11.010

29. Cao K, Wan Y, Yusufu M, Wang N (2019) Significance of Outdoor Time for Myopia Prevention: A Systematic Review and Meta-Analysis Based on Randomized Controlled Trials. Ophthalmic research:1-9. doi:10.1159/000501937

30. Deng L, Pang Y (2019) Effect of Outdoor Activities in Myopia Control: Meta-analysis of Clinical Studies. Optometry and vision science: official publication of the American Academy of Optometry 96 (4):276–282. doi:10.1097/OPX.0000000000001357

31. He M, Xiang F, Zeng Y, Mai J, Chen Q, Zhang J, Smith W, Rose K, Morgan IG (2015) Effect of Time Spent Outdoors at School on the Development of Myopia Among Children in China: A Randomized Clinical Trial. Jama 314 (11):1142–1148. doi:10.1001/jama.2015.10803

32. Rose KA, Morgan IG, Ip J, Kifley A, Huynh S, Smith W, Mitchell P (2008) Outdoor activity reduces the prevalence of myopia in children. Ophthalmology 115 (8):1279–1285. doi:10.1016/j.ophtha.2007.12.019

33. Gong Q, Janowski M, Luo M, Wei H, Chen B, Yang G, Liu L (2017) Efficacy and Adverse Effects of Atropine in Childhood Myopia: A Meta-analysis. JAMA ophthalmology 135 (6):624–630. doi:10.1001/jamaophthalmol.2017.1091

34. Lipson MJ, Brooks MM, Koffler BH (2018) The Role of Orthokeratology in Myopia Control: A Review. Eye & contact lens 44 (4):224–230. doi:10.1097/ICL.0000000000000520

35. Ostberg O, Horie Y, Feng Y (1992) On the merits of ancient Chinese eye acupressure practices. Applied ergonomics 23 (5):343–348. doi:10.1016/0003-6870(92)90296-8

36. Jadad AR, Moore RA, Carroll D, Jenkinson C, Reynolds DJ, Gavaghan DJ, McQuay HJ (1996) Assessing the quality of reports of randomized clinical trials: is blinding necessary? Controlled clinical trials 17 (1):1–12

37. Wells GA, Shea B, O’Connell D, Peterson J, Welch V, Losos M (2011) The Newcastle-Ottawa Scale (NOS) for assessing the quality of non-randomised studies in meta-analyses.. Ottawa Hospital Research Institute http://www.ohri.ca/programs/clical_epidemiology/oxford.asp Accessed 01 October 2019. Accessed 11 October 2019

38. Higgins JPT GSe (2011) Cochrane handbook for systematic reviews of interventions version 5.1.0 [updated March 2011] (available from www.cochrane-handbook.org). Cochrane Collaboration.

39. Savovic J, Weeks L, Sterne JA, Turner L, Altman DG, Moher D, Higgins JP (2014) Evaluation of the Cochrane Collaboration’s tool for assessing the risk of bias in randomized trials: focus groups, online survey, proposed recommendations and their implementation. Systematic reviews 3:37. doi:10.1186/2046-4053-3-37

40. Obtaining standard errors from confidence intervals and P values. https://handbook-5-1.cochrane.org/chapter_7/7_7_7_2_obtaining_standard_errors_from_confidence_intervals_and.htm (Accessed 11 October 2019).

41. DerSimonian R, Laird N (1986) Meta-analysis in clinical trials. Controlled clinical trials 7 (3):177–188

42. Higgins JP, Thompson SG, Deeks JJ, Altman DG (2003) Measuring inconsistency in meta-analyses. Bmj 327 (7414):557–560

43. Higgins JP, Thompson SG (2002) Quantifying heterogeneity in a meta-analysis. Stat Med 21 (11):1539–1558

44. Mantel N, Haenszel W (1959) Statistical aspects of the analysis of data from retrospective studies of disease. J Natl Cancer Inst 22 (4):719–748

45. Galbraith RF (1988) A note on graphical presentation of estimated odds ratios from several clinical trials. Stat Med 7 (8):889–894

46. Ioannidis JP, Trikalinos TA (2007) The appropriateness of asymmetry tests for publication bias in meta-analyses: a large survey. CMAJ 176 (8):1091–1096. doi:10.1503/cmaj.060410

47. Rathod V, Desai D, Alagesan J (2011) Effect of Eye Exercises on Myopia - Randomized Controlled Study. J Pharm Biomed Sci (JPBMS) 10 (10):1–4

48. Kang MT, Li SM, Peng X, Li L, Ran A, Meng B, Sun Y, Liu LR, Li H, Millodot M, Wang N (2016) Chinese Eye Exercises and Myopia Development in School Age Children: A Nested Case-control Study. Scientific reports 6:28531. doi:10.1038/srep28531

49. Lin Z, Vasudevan B, Jhanji V, Gao TY, Wang NL, Wang Q, Wang J, Ciuffreda KJ, Liang YB (2013) Eye exercises of acupoints: their impact on refractive error and visual symptoms in Chinese urban children. BMC complementary and alternative medicine 13:306. doi:10.1186/1472-6882-13-306

50. Han X, Miao H, Huang D (2014) Investigation of junior school student myopia in high - altitude Tibetan areas in Qinghai Province. Int Eye Sci 14 (5):913–915

51. Li SM, Kang MT, Peng XX, Li SY, Wang Y, Li L, Yu J, Qiu LX, Sun YY, Liu LR, Li H, Sun X, Millodot M, Wang N (2015) Efficacy of Chinese eye exercises on reducing accommodative lag in school-aged children: a randomized controlled trial. PloS one 10 (3):e0117552. doi:10.1371/journal.pone.0117552

52. Lin Z, Vasudevan B, Fang SJ, Jhanji V, Mao GY, Han W, Gao TY, Ciuffreda KJ, Liang YB (2016) Eye exercises of acupoints: their impact on myopia and visual symptoms in Chinese rural children. BMC complementary and alternative medicine 16:349. doi:10.1186/s12906-016-1289-4

53. Wang L, Du M, Yi H, Duan S, Guo W, Qin P, Hao Z, Sun J (2017) Prevalence of and Factors Associated with Myopia in Inner Mongolia Medical Students in China, a cross-sectional study. BMC ophthalmology 17 (1):52. doi:10.1186/s12886-017-0446-y

54. Huang L, Kawasaki H, Liu Y, Wang Z (2019) The prevalence of myopia and the factors associated with it among university students in Nanjing: A cross-sectional study. Medicine 98 (10):e14777. doi:10.1097/MD.0000000000014777

55. Khan A, Fahl Mar K, Brown WA (2018) The Impact of Underpowered Studies on Clinical Trial Results. The American journal of psychiatry 175 (2):188. doi:10.1176/appi.ajp.2017.17091016

56. Lu Z, Ouyang M, Zhang R, Tang X, Zhong H (2019) Association between Chinese eye exercises and onset of myopia: a meta-analysis. Int J Clin Exp Med 12 (5):4580–4588

57. Field T (2009) Massage therapy, acupressure, and reflexology. Paper presented at the Complementary and Alternative Therapies Research Washington D.C.,

58. Xiong R (2001) Survey on the nonstandard performance of Chinese eye exercises in children. Chinese Journal of School Health 22 (1)

59. Zhao R, Zhu J (2012) Beliefs and behavior related to Chinese students eye exercises among primary and secondary school teachers and students in Shanghai. Chinese Journal of School Health 33 (3)

